# VPNet: A Unified Explainable Classification Framework Using Probabilistic Circuits and Foundation Models

**DOI:** 10.1101/2025.09.30.25336962

**Authors:** Taishi Kusumoto

## Abstract

Deep learning-based classification models can learn and extract complex representations or relationships between classes. If the decision process of such AI models can be visualized in a way that humans can understand, this may create the potential for AI tools to assist in new scientific discoveries and improve trust in AI models. However, visualizations produced by current post-hoc XAI methods, including Grad-CAM and attention maps, remain theoretically questionable.

To overcome this limitation, we propose a novel theoretically supported explainable AI model for classification tasks that can handle multimodal information. Unlike conventional black-box classifiers based solely on convolutional neural networks or transformers, our approach combines probabilistic circuits with two vision transformers, DINO and CLIP, enabling probabilistic interpretability at the patch-associated embedding level, encoder level, and modality level. We demonstrate that our proposed method can pass the randomized test for saliency checks, a test that widely used post-hoc XAI methods often fail. Additionally, this study demonstrates strong generalization performance and representational capability across various classification tasks while preserving the model’s explainable structure.

## 1 Introduction

The recent advancement in deep learning techniques has raised the expectation that AI models may contribute to new scientific discoveries, potentially enabling breakthroughs across many scientific fields. From this perspective, one of the most promising deep learning techniques may be supervised classification. With an appropriately labeled dataset, if a classification model is well optimized, then visualizing its decision process may enable scientists to uncover new scientific relationships between the classes because deep learning models can recognize and learn complex features and relationships not understandable for humans.

However, traditional classification algorithms based on convolutional neural networks (CNNs)[1] or transformers[2] often complicate such visualization because of their black-box characteristics. Their complex mathematical operations, both linear and nonlinear, intricately mix the decision process used for classification, making it difficult to interpret.

To overcome this situation, several explainable AI (XAI) methods for classification have been proposed to extract the model’s decision process [3], including Grad-CAM based approaches [4], LRP based approaches [5], attention map based approaches [6], and perturbation-based approaches such as LIME [7] and SHAP [8].

However, as some studies suggest [9–12], these post-hoc XAI techniques may merely project complex nonlinear features in neural networks into a human-interpretable form, without theoretical evidence that the resulting visualization directly corresponds to the actual decision-making process of the model.

To address this limitation of such widely used post-hoc XAI techniques, we decided to take a completely different approach to visualization. Instead of extraction of the mathematical components inside CNNs or transformers, we adopted a tractable neural network, namely a probabilistic circuit [13– 16] and designed an explainable framework utilizing the interpretable structures. Using probabilistic circuits, it is possible to design tractable classification models with tractable probabilistic semantics in the decision layer. However, probabilistic circuits do not have a strong ability to extract complex feature representations compared to CNNs [1] or transformers[2], which complicates the optimization of tractable models based on probabilistic circuits.

The emergence of recent foundation models, trained with enormous computational costs and vast amounts of data, and exhibiting highly sophisticated feature extraction abilities, can help resolve this limitation. We propose a new framework that connects embedding vectors from a foundation model to probabilistic circuits without disturbing tractability by strictly preserving decomposability and smoothness [13] in the circuit. In particular, we connect embedding vectors to disjoint variable scopes so that the circuit structure remains decomposable and smooth while incorporating rich representations and domain-specific knowledge from the foundation model. With this algorithm, probabilistic circuits can leverage the strong feature extraction ability of the foundation model, enabling more complex classification tasks.

This framework can be applied to classification tasks in any domain with an appropriate foundation model, including text, images, genes, audio, and signals. However, in this paper, as one example of our framework, we propose VPNet, a 2D or 3D multimodal image classification algorithm. VPNet combines two parameter-frozen vision transformers[6], CLIP [17] and DINO[18], which are foundation models trained on large-scale image datasets, with probabilistic circuits.

In this study, our main contributions are as follows:

- We propose VPNet, a classification framework that combines frozen vision transformers with probabilistic circuits, enabling probabilistic visualization of the model’s decision process.
- We demonstrate the representational capability and generalization performance of VPNet across multiple image classification tasks, including brain tumor classification using BRISC2025 [19], pneumonia classification using ChestX-ray14 [20], and melanoma classification using dermoscopic images from the ISIC archive [21, 22].
- We demonstrate probabilistic interpretability by visualizing saliency maps and evaluating their consistency with actual defects in the MVTec anomaly detection dataset [23].

One example visualization for the melanoma classification task is shown in Figure 1. VPNet visualizes patch-associated probabilistic contributions that are directly involved in the classification decision. This differs from traditional post-hoc XAI methods, such as Grad-CAM or attention maps, whose saliency maps are computed after model training. We expect that our framework may reduce the need for traditional post-hoc XAI methods in both practice and research.

**Figure 1.**
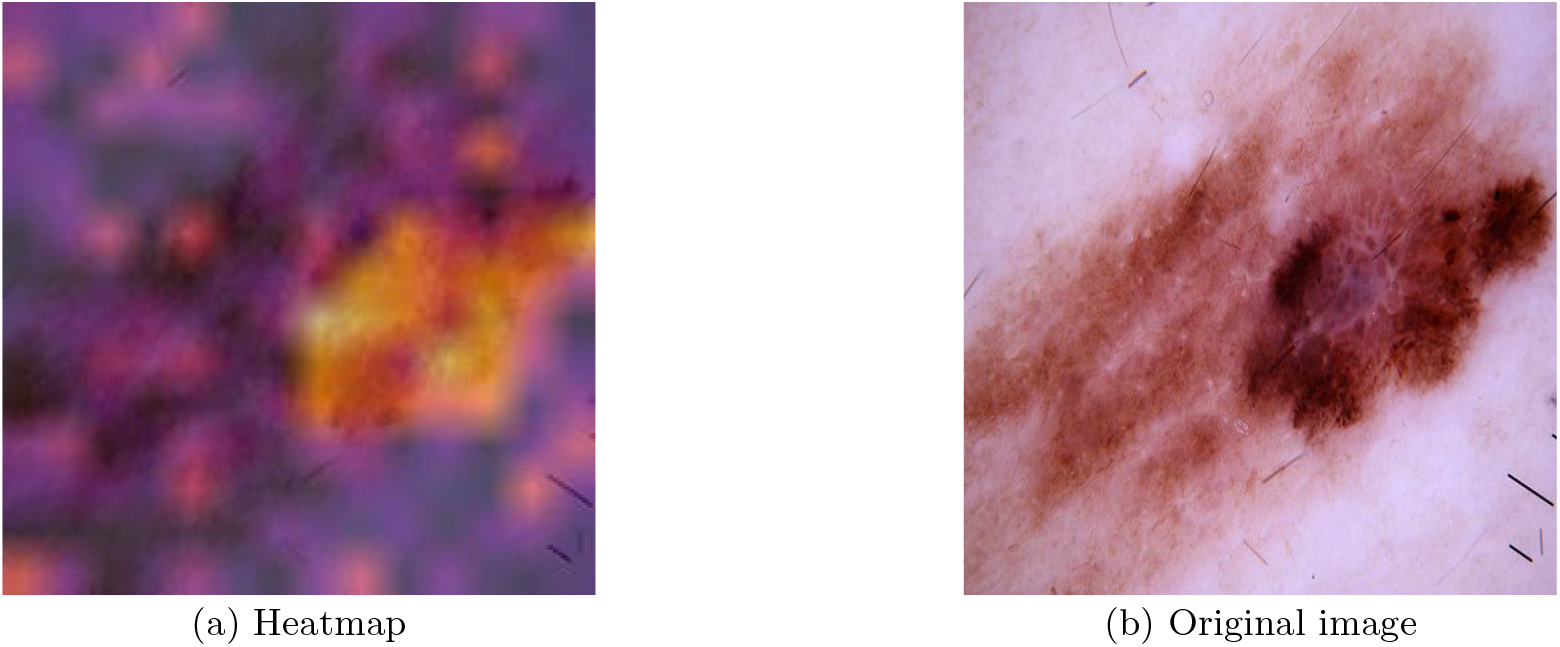
Example visualization for melanoma classification. The heatmap shows patch-associated probabilistic contributions toward the melanoma class. Brighter regions indicate higher probabilistic contributions, whereas darker regions indicate lower contributions.

## 2 Methods

### 2.1 Tractable Probabilistic Circuits

A probabilistic circuit [13] is a neural network whose outputs represent the joint probability distribution over all input scopes, provided that the circuit satisfies the following conditions: (a) the input of each leaf node corresponds to a single random variable, and (b) smoothness and decomposability are preserved.

Each image input is converted into several patch embedding vectors from a parameter-frozen vision transformer as follows:

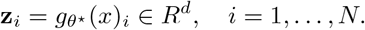

Then, each scalar of a patch vector is directly connected to each leaf node of the circuit. Each leaf node encodes a univariate Gaussian probability density function:

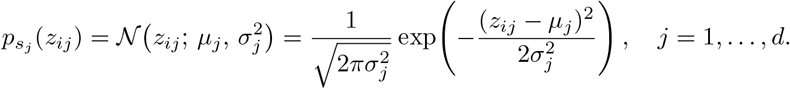

where *z*_*ij*_ denotes the *j*-th scalar of the *i*-th patch embedding **z**_*i*_.

Each scalar derived from a patch embedding vector is treated as an observed value of a random variable with a disjoint scope. This guarantees that each input unit of a probabilistic circuit has a disjoint scope.

A product node and a sum node are defined as follows:

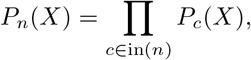

for a product node *n*, and

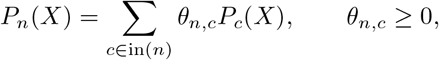

for a sum node *n*. Here, in a circuit, if all sum nodes are connected to nodes with the same scope, this preserves the smoothness, and if all product nodes are connected to nodes with disjoint scopes with each other, this preserves the decomposability. In this case, a probabilistic circuit encodes a joint distribution of all the input random variables. Our probabilitic circuit encodes the joint probabilistic distribution of all the input scaler components, which can be assumed as the same to the joint probabilitic distribution of the patch embedding vector written as,

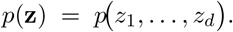

For memory efficiency and to accommodate variable-length inputs, we adopted a **RAT-SPN– based** algorithm [24], in which a compact probabilistic circuit, receiving a single patch embedding vector as input, is shared across all patch embeddings within an image and a ViT. We assume that each patch embedding vector is a subset of different random variables; in other words, the patch embedding vectors have disjoint scopes with each other. If a PC such as *C*_*B*_ has the same DAG as the computational graph of *C*_*A*_ and parameters *θ*_*B*_ = *θ*_*A*_, but the input has a disjoint scope from the input of *C*_*A*_, the two circuits encode different functions with disjoint scopes. For memory efficiency and to accommodate variable-length inputs, we adopted a **RAT-SPN–based** algorithm [24], in which a compact probabilistic circuit, receiving a single patch embedding vector as input, is shared across all patch embeddings within an image and a ViT. We assume that each patch embedding vector is assigned to a distinct variable scope, and that the scopes of different patch embedding vectors are mutually disjoint. If a PC such as *C*_*B*_ has the same DAG as the computational graph of *C*_*A*_ and parameters *θ*_*B*_ = *θ*_*A*_, but the input has a disjoint scope from the input of *C*_*A*_, the two circuits encode different functions with disjoint scopes. This ensures that any two such distributions from shared probabilistic circuits can be integrated by product nodes as a joint probabilistic distribution:

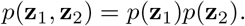

This integrated joint probabilistic distribution has a disjoint scope *S*_1_ ∪ *S*_2_. By repeating this process, a joint distribution with an integrated scope *S*_1_ ∪ *S*_2_ ∪ *· · ·* ∪ *S*_*N*_ is obtained:

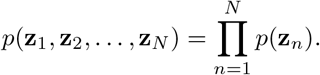

To increase the representational capability, we prepared separate probabilistic circuits for different encoders and modalities, with the same DAG but different parameters. Each embedding vector is assigned a disjoint scope. This assignment allows outputs to be integrated through product nodes while preserving decomposability. For optimization, the output distributions associated with all patch embeddings across every encoder and modality are integrated into a joint distribution using product nodes:

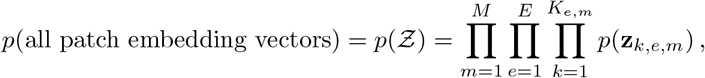

where

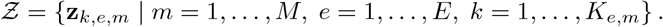

Here, **z**_*k,e,m*_ denotes the *k*-th patch embedding vector generated by encoder *e* from modality *m*, and *K*_*e,m*_ denotes the number of patch embedding vectors generated for the corresponding encoder and modality pair.

For classification, each class has its own joint probability distribution defined above. The corresponding class-conditional joint distribution is written as

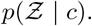

Here, a PC associated with class *c* is not connected to any PC associated with another class, and its parameters are optimized independently of those of the PCs associated with the other classes.

In summary, the number of probabilistic circuits required is determined as follows:

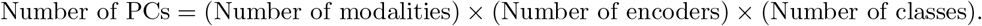

In this paper, for binary 2D image classification, we use two frozen ViT encoders, DINO and CLIP, with a single modality. Therefore, four PCs are required.

The overview of this architecture for two modalities, two encoders, and two classes is shown in Figure 2

**Figure 2.**
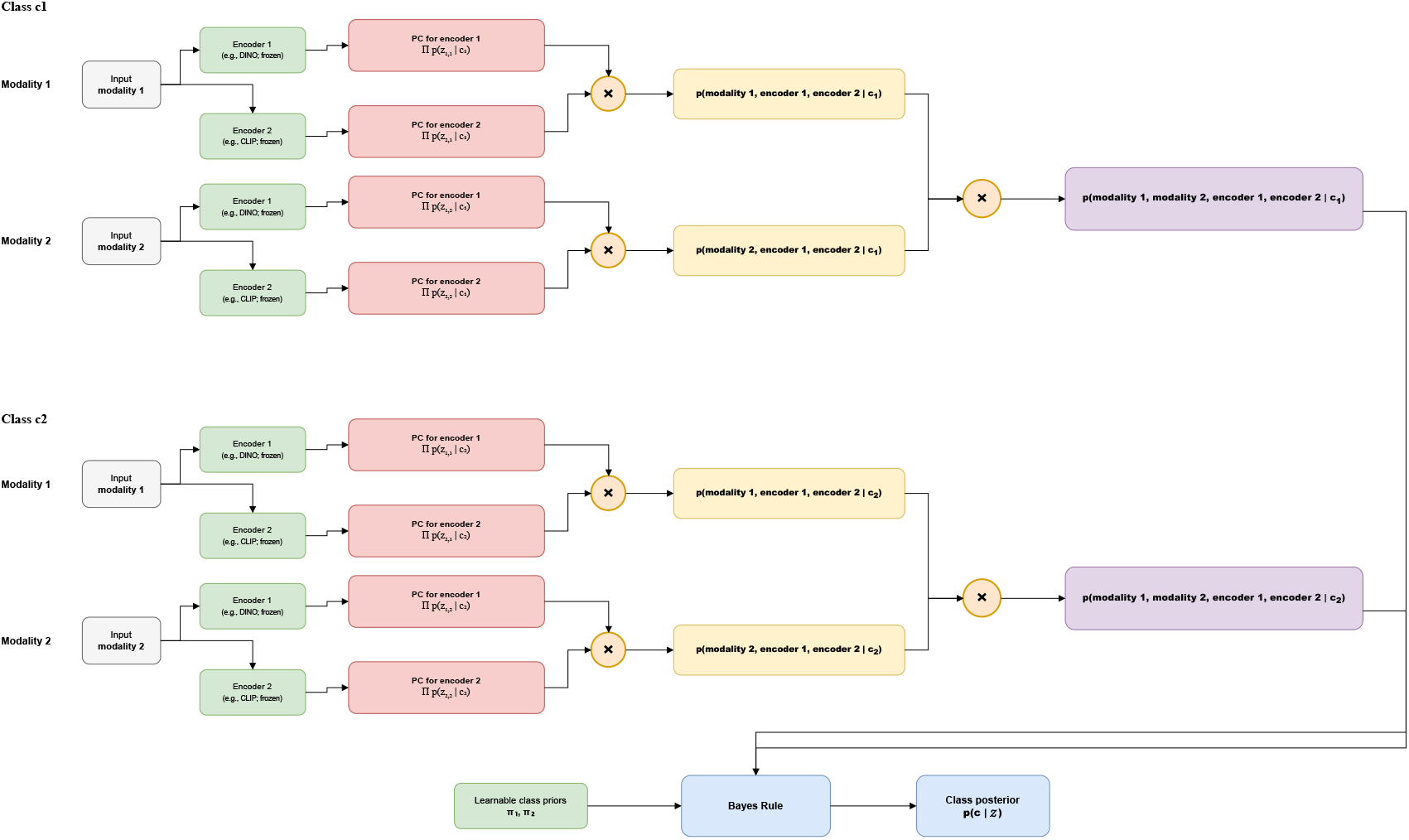
Overview of the VPNet architecture for two modalities, two encoders, and two classes.

### 2.2 Optimization for classification

Here, apart from the probabilistic circuits, we set an additional simple and learnable network. This network represents a categorical distribution of the class prior, defined as

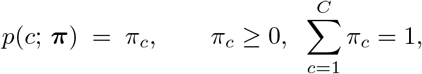

where ***π*** = (*π*_1_, … , *π*_*C*_) denotes the learnable class prior parameters.

The joint distribution factorizes by the product rule as

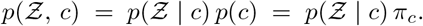

The class posterior is then given by Bayes’ rule:

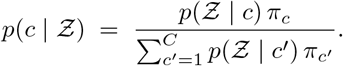

This class posterior is used for the optimization process. As loss functions, we define the cross-entropy loss [25],

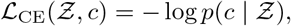

and the Shannon entropy loss to avoid overfitting [26],

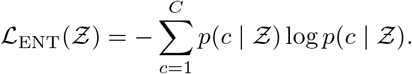

In this paper, we adopt the following fractional likelihood [27]in order to stabilize training. For class *c*, we define

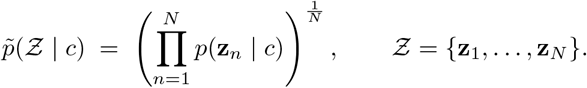

Then, we compute a fractional posterior by applying Bayes’ rule to the geometric-mean likelihood:

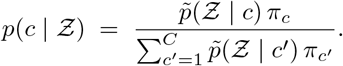

Although this differs from the exact Bayesian posterior based on the full product likelihood, the relative ordering of the likelihoods is preserved, thereby not affecting the interpretability of the model.

### 2.3 Extraction of the Model’s Decision-Making Process via Marginal Class-Conditional Distributions

During inference, we obtain the class-conditional likelihood for any chosen information, including a *region* (a set of patch indices), *modality*, or *encoder*, by simply extracting the corresponding likelihood through a forward pass and factorization of the corresponding root nodes.

We denote the returned class-conditional likelihood by

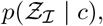

where ℛ ⊆{1, … , *N*} denotes the set of indices corresponding to the focused information. The resulting joint distribution is equivalent to the marginal distribution obtained when all other inputs are unobserved.

For the focused information, which class receives a greater probabilistic contribution from the model can be quantified by subtracting the class-conditional likelihoods of the respective classes. For instance, in binary classification, we define

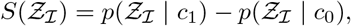

as the probabilistic contribution score for class *c*_1_ from the focused information. A positive value indicates stronger evidence for class *c*_1_, whereas a negative value indicates stronger evidence for class *c*_0_.

The probabilistic contributions of different information sources can also be compared using their probabilistic contribution scores, 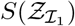 and 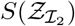. For the construction of saliency maps, we use the patch-level probabilistic contribution score *S*(*Ƶ*_{*n*}_) for each patch *n*.

### 2.4 Datasets for 2D image classification

We prepared a brain tumor classification dataset from MRI images, BRISC2025[19], and a pneumonia classification dataset from chest X-ray images, ChestX-ray8[20], to demonstrate the tractability and classification ability of our model.

BRISC2025 has 4 specific classes, including no tumor, glioma, meningioma, and pituitary, with 1067, 1147, 1329, and 1457 training images, and 140, 254, 306, and 300 validation images, respectively. We integrated all the training and validation images except the no-tumor class into a single tumor class. Then, the training samples in the no-tumor class were oversampled so that the ratio of the two classes is nearly equal. This dataset has ground truth segmentation masks corresponding to tumor regions for every image in the tumor class.

We used ISIC 2018 challenge archive.[22] We converted the task into a binary classification problem by defining melanoma (mel) as the positive class and merging all remaining categories (akiec, bcc, bkl, df, nv, and vasc) into a single negative class. Training, validation, and test datasets are used without any modification. The original dataset splits were used without modification: the training set contained 10,015 images (1,113 melanoma and 8,902 others), the validation set contained 193 images (21 melanoma and 172 others), and the test set contained 1,512 images (171 melanoma and 1,341 others).

ChestX-ray8 comprises 2 specific classes, normal and pneumonia, with 1341 and 3875 training images, 8 and 8 validation images, and 234 and 350 test images, respectively. For this dataset, we did not make any modifications and use it directly.

In addition, we prepared a simpler dataset for anomaly detection, the MVTec dataset.[23] We used two of its categories, zipper and tile, comprising two specific classes, good and NG.

Mvtec-zipper includes 240 training images for the good class, and 32 and 119 test images for the good and NG classes, respectively. We selected the first 21 images from the NG test set and used them as training images for the NG class. The training NG images were oversampled so that the ratio of the two classes was nearly equal. All the remaining test images were used as validation images.

MVTec Tile contains 230 training images for the good class, and 33 and 84 test images for the good and NG classes, respectively. We selected 26 images and 5 images from the NG test dataset and 5 images from the good training dataset, and used them as training images and validation images, respectively.

### 2.5 Experimental setting for 2D image classification

In the experiments, the probabilistic circuits were initialized with the following default configurations: for ChestX-ray8, a depth of 5, 64 latent variables, 3 repetitions, and 2 mixture pieces per leaf were used; for MVTec Tile, a depth of 2, 32 latent variables, 2 repetitions, and 4 mixture pieces per leaf were used; and for the remaining datasets, a depth of 2, 16 latent variables, 2 repetitions, and 2 mixture pieces per leaf were used. Two frozen vision transformers, DINO ViT-B/16 and CLIP ViT-B/16, were used as the backbones of the circuits. Since these ViTs are optimized for 224 × 224 images, all input images were resized to this resolution. As a result, each input image was divided into 196 patches. The Adam optimizer [28] was adopted for training.

During training, we set a threshold of 0.5 for the anomaly class, then calculated the accuracy on the validation dataset, and saved the parameters that achieved the maximum accuracy. Gaussian, speckle, and salt-and-pepper noise are applied to the input images as part of data augmentation.

## 3 Results

### 3.1 Overall Classification Performance for 2D image classification

Table 1 shows the AUROC, Average Precision (AP), and maximum accuracy from the experimental datasets.

**Table 1:** Test metrics for each dataset. Values are AUROC, Average Precision (AP), and Max Accuracy with the threshold used.

| Dataset | AUROC | AP | Max Acc. @ thr |
| --- | --- | --- | --- |
| BRISC2025 (2-class) | 0.994 | 0.9999 | 0.972 @ 0.3860 |
| MVTec (tile) | 0.9571 | 0.9766 | 0.9070 @ 0.1824 |
| ChestX-ray8 | 0.9183 | 0.9385 | 0.8667 @ 0.8680 |
| ISIC archive | 0.8705 | 0.5493 | 0.9113 @ 0.6257 |

### 3.2 Patch-level Probabilistic Contribution for 2D image classification

#### 3.2.1 Posterior-based visualization

Figure 3 illustrates two inferno heatmaps [29] based on the class posteriors for several anomaly images in the test dataset of MVTec Tile, together with the corresponding original images. It is observed that both the dark purple regions in the heatmap from the negative class and the dark red regions in the heatmap from the positive class cover the defective parts in the originals. This indicates that the basis of the model’s decision concentrates on the defective areas.

**Figure 3.**
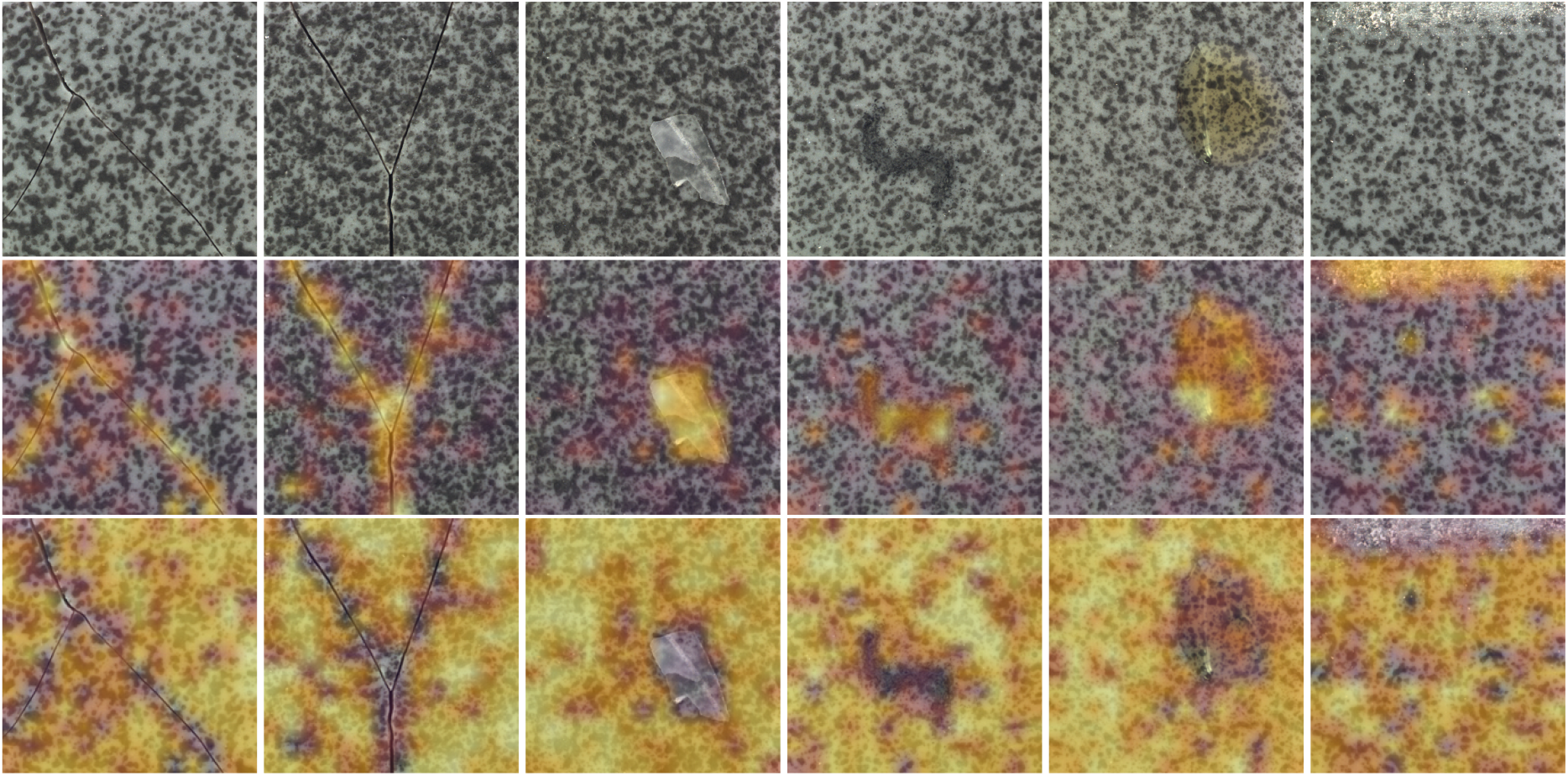
Three rows (top to bottom): original, heatmap (NG), heatmap (good). Six columns show different samples in MVTec-tile.

#### 3.2.2 Likelihood-based visualization

Instead of class posteriors, Figure 4 from the test set of the ISIC archive and Figure 5 from BRISC 2025 integrate the patch-level probabilistic contributions into a single heatmap using the patch-wise class-likelihood difference defined in Section 2.5.

**Figure 4.**
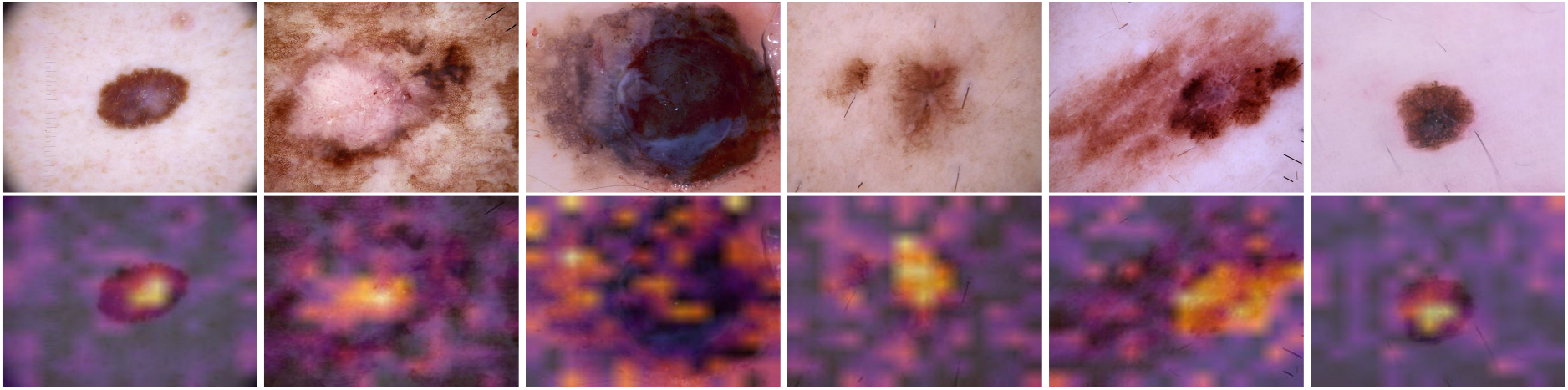
Likelihood-based heatmaps on the ISIC test set. The top row shows the original images, and the bottom row shows the corresponding heatmaps based on patch-wise likelihood differences. Brighter regions indicate stronger contributions toward the melanoma class.

**Figure 5.**
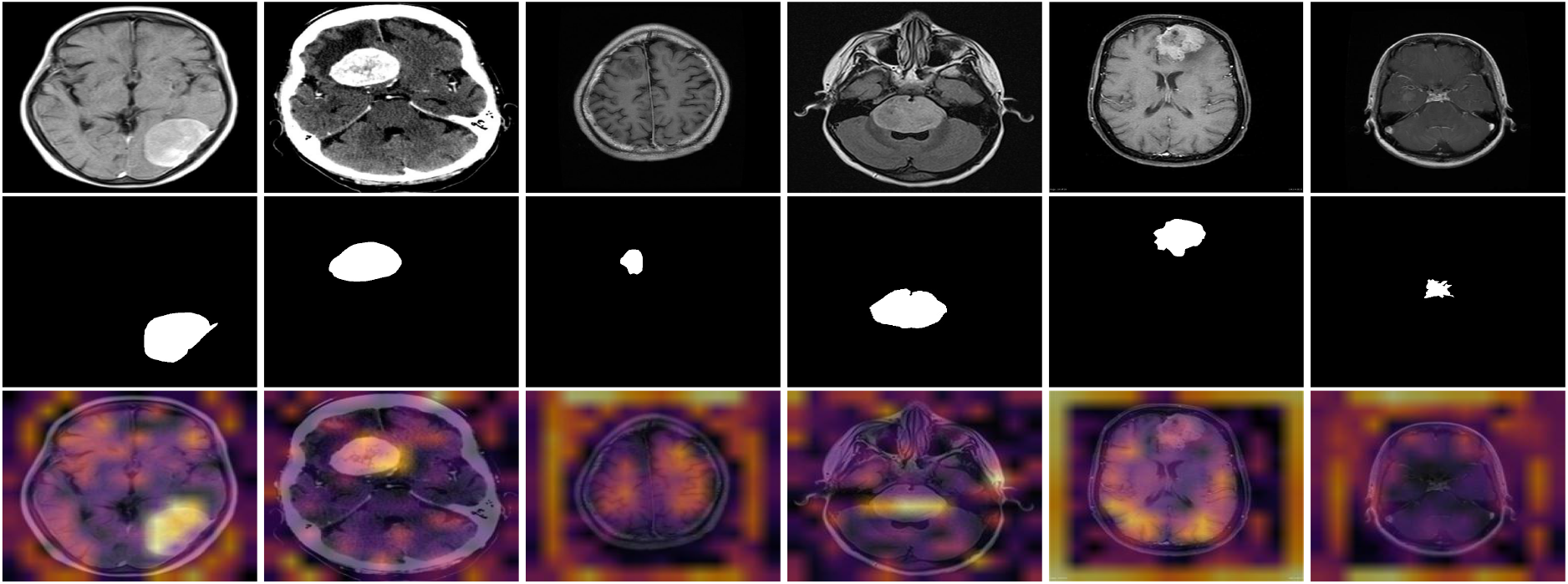
Likelihood-based heatmaps on the BRISC 2025 dataset. The top row shows the original images, the middle row shows the ground-truth annotations, and the bottom row shows the corresponding heatmaps based on patch-wise likelihood differences.

In Figure 4, brighter regions are observed to concentrate on the lesion areas of the images, whereas some regions within the lesions are highlighted by darker colors. This indicates that the model distinguishes between more melanoma-like and less melanoma-like regions within lesions and assigns corresponding probabilistic contributions.

On the other hand, some heatmaps from the test set of the ISIC archive highlight tumor regions with strong bright colors, whereas others strongly highlight the background, as shown in Figure 5. This suggests that the model adapts its focus depending on the input image.

#### 3.2.3 Randomized test

We checked whether the heatmaps were obtained during the training process by comparing them with heatmaps from a model with randomly initialized probabilistic circuits. Figure 6 presents the differences in the heatmaps after training for likelihood from both DINO and CLIP, DINO only, and CLIP only, respectively. This figure supports the following findings: (a) the heatmaps of the classification model are obtained through optimization and do not originate from the attention maps of the vision transformers, (b) the DINO circuits and the CLIP circuits learn different decisions independently during training, and the representations they capture are fundamentally different.

**Figure 6.**
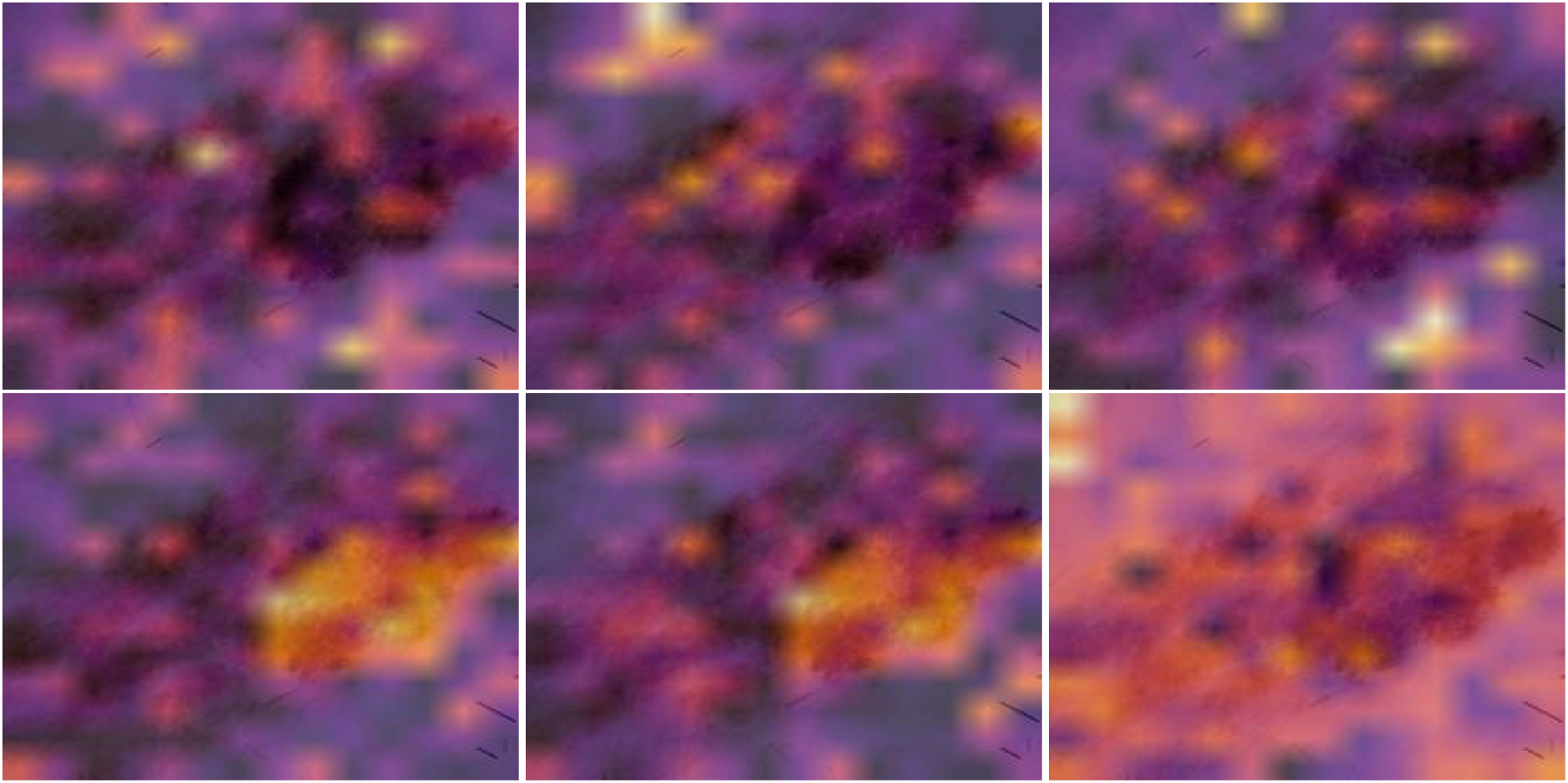
Comparison of likelihood-based heatmaps from randomly initialized and trained probabilistic circuits. The top row shows heatmaps obtained from randomly initialized circuits, while the bottom row shows those obtained after training. From left to right, the columns correspond to DINO+CLIP, DINO, and CLIP, respectively.

For quantitative analysis, we computed Spearman’s rank correlation, SSIM, and HOG-based Pearson correlation between heatmaps from randomly initialized PCs and those from trained DINO, CLIP, or DINO+CLIP PCs for all models using the test datasets. The results are summarized in Table 2. Across all results, the mean Spearman rank correlations ranged from *−*0.120 to 0.158, while the mean HOG-based Pearson correlations ranged from 0.067 to 0.108, suggesting little correlation between the heatmaps. Although the mean SSIM values were slightly higher, they remained between 0.364 and 0.462. Overall, these results do not support the hypothesis that the heatmaps produced by the trained models depend on those produced by the randomly initialized models.

**Table 2:** Quantitative comparison between heatmaps obtained from randomly initialized and trained probabilistic circuits.

| Dataset | Encoder | Spearman rank correlation |  |  | SSIM |  |  | HOG Pearson correlation |  |  |
| --- | --- | --- | --- | --- | --- | --- | --- | --- | --- | --- |
|  |  | Min | Max | Mean | Min | Max | Mean | Min | Max | Mean |
| BRISC2025 | CLIP | -0.4303 | 0.1802 | -0.1136 | 0.2103 | 0.5615 | 0.3648 | -0.0028 | 0.1562 | 0.0730 |
|  | DINO | -0.5663 | 0.4640 | -0.1092 | 0.2089 | 0.5668 | 0.3795 | -0.0013 | 0.1657 | 0.0776 |
|  | DINO+CLIP | -0.5644 | 0.4617 | -0.1199 | 0.2176 | 0.5363 | 0.3722 | -0.0186 | 0.1493 | 0.0672 |
| ISIC 2018 Melanoma | CLIP | -0.3072 | 0.3089 | 0.0254 | 0.2864 | 0.5105 | 0.3923 | 0.0224 | 0.1458 | 0.0753 |
|  | DINO | -0.2000 | 0.4261 | 0.0766 | 0.3223 | 0.5520 | 0.4249 | 0.0254 | 0.1495 | 0.0766 |
|  | DINO+CLIP | -0.2076 | 0.2876 | 0.0422 | 0.2784 | 0.5384 | 0.4045 | 0.0184 | 0.1448 | 0.0742 |
| MVTec Tile | CLIP | -0.0780 | 0.3081 | 0.1169 | 0.2927 | 0.5389 | 0.4606 | 0.0404 | 0.1753 | 0.1079 |
|  | DINO | -0.2107 | 0.3013 | 0.0518 | 0.2574 | 0.5150 | 0.3776 | 0.0130 | 0.1329 | 0.0720 |
|  | DINO+CLIP | -0.1539 | 0.3178 | 0.0792 | 0.2525 | 0.5997 | 0.4299 | 0.0211 | 0.1567 | 0.0885 |
| ChestX-ray8 | CLIP | -0.1390 | 0.4638 | 0.1582 | 0.2934 | 0.5799 | 0.4622 | 0.0094 | 0.1465 | 0.0758 |
|  | DINO | -0.3948 | 0.4672 | -0.0790 | 0.2430 | 0.5080 | 0.3702 | 0.0171 | 0.1463 | 0.0796 |
|  | DINO+CLIP | -0.3496 | 0.2856 | -0.0344 | 0.2521 | 0.4934 | 0.3636 | 0.0015 | 0.1333 | 0.0719 |

The results are summarized in Table 2. Across all results, the mean Spearman rank correlations ranged from *−*0.120 to 0.158, while the mean HOG-based Pearson correlations ranged from 0.067 to 0.108, suggesting little correlation between the heatmaps. Although the mean SSIM values were slightly higher, they remained between 0.364 and 0.462. Overall, these results do not support the hypothesis that the heatmaps produced by the trained models depend on those produced by the randomly initialized models.

## 4 Discussion and Conclusion

### 4.1 Comparison with other XAI models

As some studies indicate, widely used post-hoc XAI methods, such as gradient- and backpropagation-based approaches, including Grad-CAM [4] and LRP [30, 31], attention-based approaches [32, 33], and perturbation-based approaches such as LIME [7] and SHAP [8], do not have solid theoretical support regarding whether their saliency maps truly represent the model’s decision process due to the black-box nature of the underlying models.[9–12]

First, for attention-map-based methods and backpropagation-based approaches, neither the visualization of internal feature representations as a map nor the amount of propagated gradients generally guarantees that relative differences across regions represent their relative importance to the model’s decision.

Second, for perturbation-based approaches, even if the contribution to the probabilistic score of a masked feature can be computed in a theoretically supported way, the effect of masking the information on the score may reflect the model’s sensitivity or robustness to the chosen perturbation, which does not guarantee that the relative magnitude of the effect corresponds to the relative importance assigned by the model in making its decision.

Additionally, it is well known that such backpropagation-based methods often fail the randomized test; when the model parameters are randomly initialized, the saliency maps remain largely unchanged, suggesting that the saliency maps are a product of the feature maps rather than an outcome of the training process.[10]

On the other hand, our proposed model is not on the same playing field as these methods, as our approach does not aim to make a black-box model explainable. Instead, the explainability of our approach is theoretically supported. A model with an interpretable structure is optimized for a specific task. The saliency maps directly reflect the model’s decision process, and each decomposed probabilistic contribution at the embedding-vector, encoder, and modality levels can be computed as a marginal probabilistic distribution when all other inputs are treated as unobserved.

Additionally, as described in Section 3.2.3, our model can overcome the randomized test, experimentally supporting that the saliency maps produced by our model are an outcome of the optimization process. Furthermore, by combining foundation models with strong representational ability and probabilistic circuits, our model compensates for one of the main weaknesses of probabilistic circuits, namely their limited representational capability. This is supported by the high classification performance across various datasets, including complex tasks such as melanoma classification.

Moreover, other intrinsically interpretable models, such as Generalized Additive Models (GAMs) [34], Explainable Boosting Machines (EBMs) [35], and Neural Additive Models (NAMs) [36], do not provide a unified probabilistic framework that enables embedding-level, encoder-level, and modality-level interpretability, as VPNet does.

The advantages of VPNet compared with post-hoc XAI methods and intrinsically interpretable models are summarized in Table 3.

**Table 3:** Comparison of post-hoc XAI methods, intrinsically interpretable models, and VPNet.

| Category | Representative methods | Foundation model compatibility | Decision-faithful interpretability | Unified probabilistic interpretability |
| --- | --- | --- | --- | --- |
| Post-hoc explanation methods | Grad-CAM [4]<br>LRP [5]<br>Attention maps [6]<br>LIME [7]<br>SHAP [8] | ○ | △ | △ |
| Intrinsically interpretable models | Generalized Additive Models (GAMs) [34]<br>Explainable Boosting Machines (EBMs) [35]<br>Neural Additive Models (NAMs) [36] | △ | ○ | △ |
| VPNet | Frozen foundation models<br>+ probabilistic circuits | ○ | ○ | ○ |

○: fully supported by the model design. △: partially supported or supported only under limited conditions.

### 4.2 Importance of Theoretically Supported Explainability

Due to the lack of guaranteed faithfulness in saliency maps generated by post-hoc explanation methods, it is difficult to determine whether the observed saliency patterns originate from the model’s optimization process or from artifacts of the explanation method itself. This ambiguity is problematic for both research and practical applications. On the other hand, the explainability of VPNet can be theoretically attributed to the model’s optimization process. Classification models with high classification performance cannot automatically be considered trustworthy, because the classification of test datasets can often be solved using meaningless features due to data leakage. However, VPNet enables users to assess whether the optimized model relies on plausible or spurious features by inspecting its saliency maps. In addition, because deep-learning models can capture and learn complex relationships and rules that humans may not recognize when solving a given task, VPNet has the potential to serve as a tool for new scientific discovery by utilizing the representational capacity of domain-specific foundation models and visualizing its decision process in a human-understandable manner.

### 4.3 Scope of Theoretically Supported Explainability

As described above, probabilistic contributions from VPNet are tractable at the embedding-vector level. However, this does not automatically guarantee patch-level explainability, because a patch embedding vector generated by a frozen ViT is obtained through nonlinear mathematical operations that may incorporate information from other patches. Therefore, the patch-level interpretation is valid only to the extent that each patch embedding vector remains representative of the corresponding image patch. The representativeness of patch embeddings is not theoretically guaranteed for either DINO [18] or CLIP [17], because these models are not explicitly trained to ensure that each patch embedding remains locally representative of its corresponding image patch. Therefore, for theoretically guaranteed regional explainability, distinct inputs are required. For example, if each image region is encoded independently without information exchange across regions, the probabilistic contribution of each embedding corresponds to the regional probabilistic contribution for the classification task. Additionally, the decomposable probabilistic contributions at the modality and encoder levels described in this study are theoretically explainable because these inputs are generated independently by separate modalities or frozen encoders before being connected to their corresponding probabilistic circuits. After optimization, these probabilistic contributions enable us to understand which information the model learned to regard as more useful during the optimization process. During inference, they can also be selectively removed to examine their effects on classification performance and model trustworthiness. Table 4 summarizes the theoretical support for each decomposed information level in VPNet.

**Table 4:** Summary of theoretical support for each decomposed information level in VPNet.

|  | Embedding-vector level | Encoder level | Modality level |
| --- | --- | --- | --- |
| **Theoretical support** | △ | $\circ$ | $\circ$ |

△ : partial theoretical support depending on encoder-level representativeness.

### 4.4 Evaluation of Benchmarks

Although our model shows strong performance on the BRISC2025 dataset, some saliency maps highlight background regions rather than brain regions. Unless tumor patients exhibit a consistent structural bias in brain geometry, this observation suggests that the dataset may not accurately represent the intended classification task between tumor and non-tumor patients. Instead, the model may be leveraging spurious correlations, such as differences in age, sex, or imaging conditions. From this perspective, our model has the potential to serve as a tool for assessing the validity of benchmark datasets.

### 4.5 Extensibility

The functionality of our proposed method is not limited to two-dimensional image classification tasks. As demonstrated in Appendix B, the framework can be extended to three-dimensional multimodal classification tasks without compromising its theoretically supported explainable structure.Similarly, it can be readily extended to various explainable classification tasks by replacing patch-embedding vectors with embeddings from other foundation models. Indeed, we proposed DVPNet [37], a gene classification framework in which the model identifies genes that are inherently important for biological classification tasks using a biological foundation model, the Nucleotide Transformer [38].

### 4.6 Future Study

In this paper, we showed that VPNet provides theoretically supported explainability while utilizing the domain-specific representational capacity of foundation models. This contrasts with the limited theoretical support for the explainability of state-of-the-art post-hoc XAI methods. In future research, we will further explore the usefulness of VPNet through applied studies. In particular, we aim to investigate whether VPNet can reveal scientifically meaningful patterns that may not be easily recognized by humans, by analyzing its explanations in comparative studies of scientifically important entities.

## Data Availability

This study used only openly available datasets that were released to the public prior to the initiation of the study. Specifically, we used BRISC2025 (brain tumor MRI, https://arxiv.org/abs/2501.01836), ChestX-ray8 (https://nihcc.app.box.com/v/ChestXray-NIHCC), and MVTec Anomaly Detection (https://www.mvtec.com/company/research/datasets/mvtec-ad), all of which are publicly accessible for research purposes. No new human data were collected and no restricted or application-based datasets were used.

## A Learnable Parameters of the Probabilistic Circuit

### A.1 Learnable parameters of probabilistic circuit

To increase the number of learnable parameters, in other words, the representational capability of our probabilistic circuit, we adopt a **RAT–SPN-based**[24] algorithm under the PyJuice framework [39](although the PyJuice library itself was not directly used in our implementation). In our implementation, it is possible to specify an integer for each of the following hyperparameters:

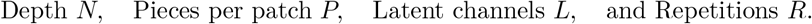

The *depth N* determines how many times the set of scalar components is recursively divided into two disjoint subsets. Each division corresponds to one layer of the RAT-SPN structure, and repeating this operation *N* times yields 2^*N*^ scopes at the leaf level. The probabilistic circuit learns the relationships between scalar components within each scope through sum nodes:

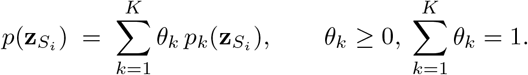

This structure ensures the **smoothness** of the probabilistic circuit, since sum nodes operate only within the same scope. Finally, the overall joint probability density of a patch vector is computed from the product node that connects the probabilities from all disjoint scopes:

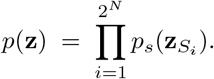

Here, **decomposability** is guaranteed because the scopes *S*_*i*_ are mutually disjoint, ensuring that each subcircuit represents a distinct factor of the full joint distribution.

The hyperparameter *Pieces per patch P* increases the number of learnable parameters at each leaf node by introducing multiple Gaussian components whose outputs are linearly combined through softmax-normalized weights. In other words, the univariate Gaussian probability density function at a leaf node is extended to include *P* parameterized Gaussian functions as follows:

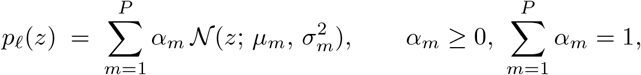

This operation simply increases the parameter capacity of the leaf node without changing the overall circuit structure.

The number of latent channels *L* determines the internal width of the probabilistic circuit at each leaf node. Each leaf node outputs an *L*-dimensional vector, in which each component represents an distinct latent subspace that models different statistical aspects of the input variable. Formally, for an input variable *z*, the leaf node produces

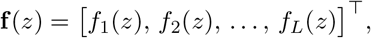

where each latent channel *f*_*l*_(*z*) has its own learnable parameters and contributes to the overall representation of the input. As all latent channels share the same scope, the outputs are integrated by sum nodes placed just above the root, which preserves the overall smoothness of the probabilistic circuit. To increase the number of learnable parameters, a linear mapping is applied to the latent outputs before the final integration as follows:

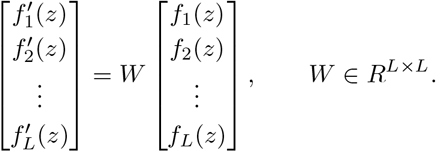

Each row of *W* corresponds to a sum node whose coefficients are normalized by a softmax function. Let the *i*-th row of *W* be denoted as

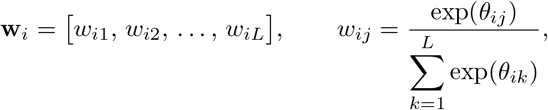

where *θ*_*ij*_ are learnable parameters. Then the output of the *i*-th sum node is

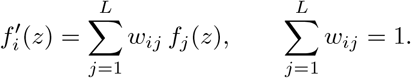

This formulation ensures that each row of *W* performs a weighted average over all latent channels, while maintaining probabilistic consistency (nonnegative coefficients summing to one).

Here, the learnable matrix *W* mixes the latent channels to enhance representational flexibility while keeping the same scope. Finally, the transformed channels are integrated by a sum node:

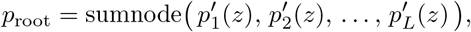

where each 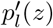 corresponds to the probability contribution from the *l*-th transformed latent channel.

Repetition *R* determines how many times the probabilistic circuit is duplicated. All duplicated PCs share an identical scope at their root nodes, so their root probabilities can be integrated by a single sum node without violating smoothness. This is represented as

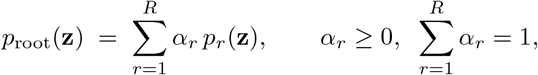

where each *p*_*r*_(**z**) denotes the probability computed by the *r*-th replicated circuit sharing the same scope, and *α*_*r*_ represents the convex weight of the root-level sum node. Because all repetitions have identical scopes, this integration by the sum node preserves the smoothness property of the circuit.

## B Brain age classification using multimodal 3D MRI images

VPNet can extend its scope to both multi-dimensional and multi-modal classification tasks beyond conventional two-dimensional and single-modal image classification without losing tractability or explainability. To demonstrate this capability, we implemented a 3D multi-modal image classification experiment. Specifically, we used the IXI 3D brain MRI dataset, which contains T1- and T2-weighted MRI volumes from 619 individuals along with patient information of age. [40]. We grouped the subjects into two age categories: *age1* (19–33 years) and *age4* (60–83 years). Any volumes outside these age ranges were excluded from the dataset. The *age1* group contained 101 training volumes, 14 validation volumes, and 30 test volumes, while the *age4* group contained 94 training volumes, 14 validation volumes, and 30 test volumes.

Each 3D MRI volume in each modality was first divided into individual 2D slices. Then, every slice was converted into multiple patch embedding vectors by two vision transformers (ViTs): DINO and CLIP. All patch vectors derived from the same 3D volume share a single probabilistic circuit. This concept is illustrated in Figure 7.

**Figure 7.**
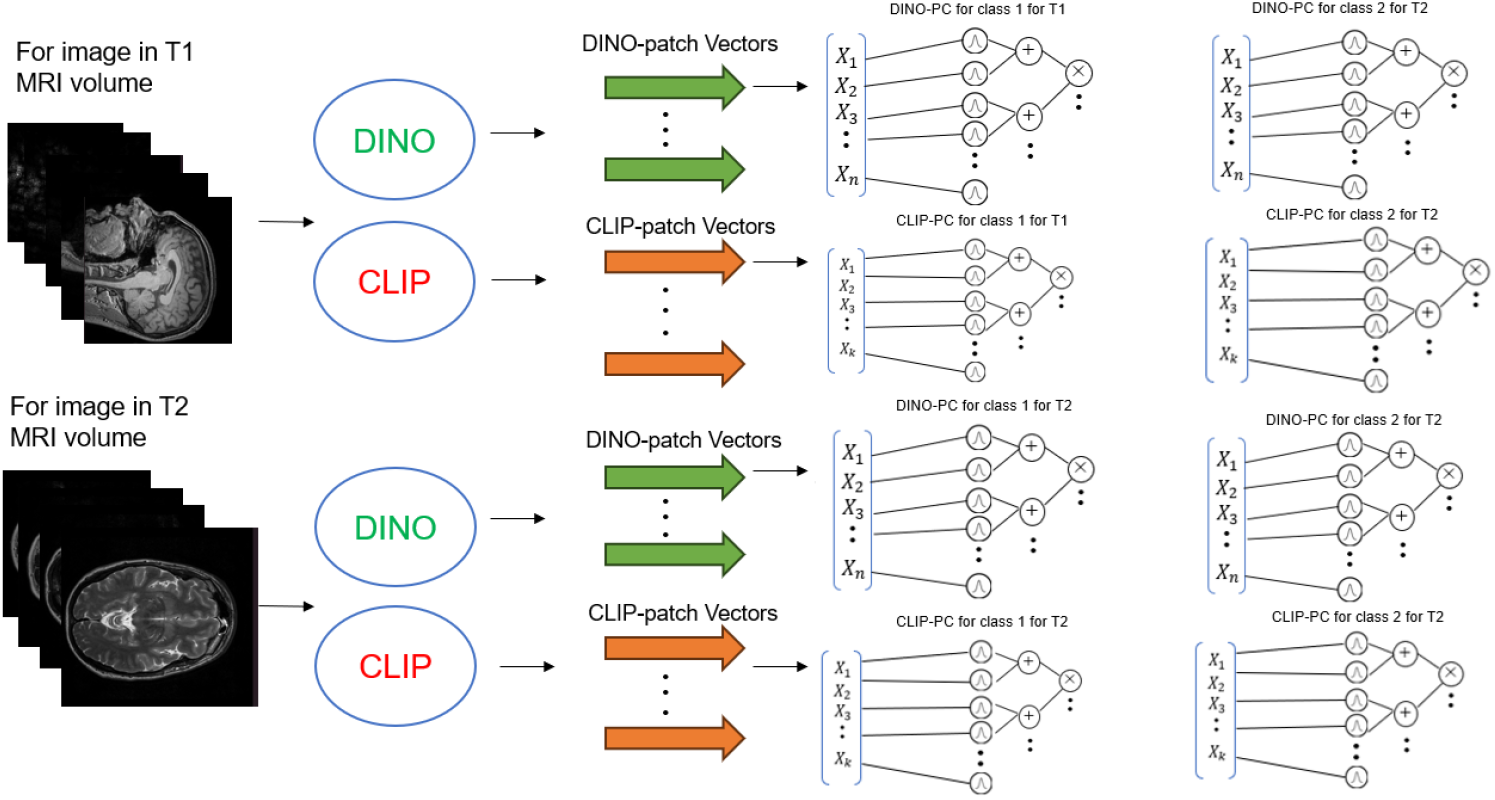
Overview of the 3D multimodal classification framework. Each 3D MRI volume is divided into slices, and each slice is transformed into patch embedding vectors by DINO and CLIP. The number of probabilistic circuits is eight, corresponding to two modalities, two encoders, and two classes.

To maintain the probabilistic interpretability, the joint posterior probability should ideally be used for optimization. However, as with most deep learning models, computing gradients through a single forward–backward pass requires storing all intermediate activations, which consumes significant memory. To address this issue, we divided the optimization into two forward passes and one backward pass.

In the first forward pass, the parameters of all probabilistic circuits were frozen, and only the joint posterior probability *P* (*c* | **Z**) was computed through all inputs *without* storing any intermediate state.Then, the overall loss was computed as the sum of the cross-entropy loss and the entropy regularization term as follows:

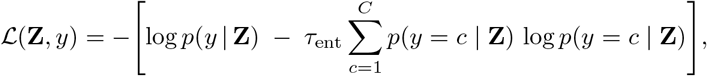

where *τ*_ent_ is a hyperparameter that controls the contribution of the entropy regularization term.

In the second forward pass, the same patch vectors were re-input into the probabilistic circuits. Based on the intermediate states of each circuit and the overall loss function defined above, the partial gradients of the loss with respect to the parameters of each probabilistic circuit were computed as follows:

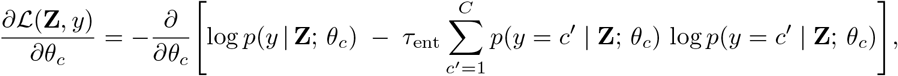

This partial gradient was computed for all the patch vectors and averaged as the overall gradient as follows:

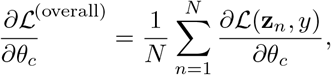

where *N* denotes the total number of patch embedding vectors in the case. Finally, the parameters of the probabilistic circuits were updated based on this overall gradient.

This process was repeated for all the samples within an epoch.

### B.1 Overall Classification Performance for 3D multimodal image classification

We optimized this model for 30 epochs with the following hyperparameters: a depth of 5, 64 latent variables, 3 repetitions, and 2 mixture pieces per leaf. Figure 8 displays the confusion matrices for the training, validation, and test samples, based on the posterior probabilities computed from the three input settings: T1+T2, T1 only, and T2 only.

**Figure 8.**
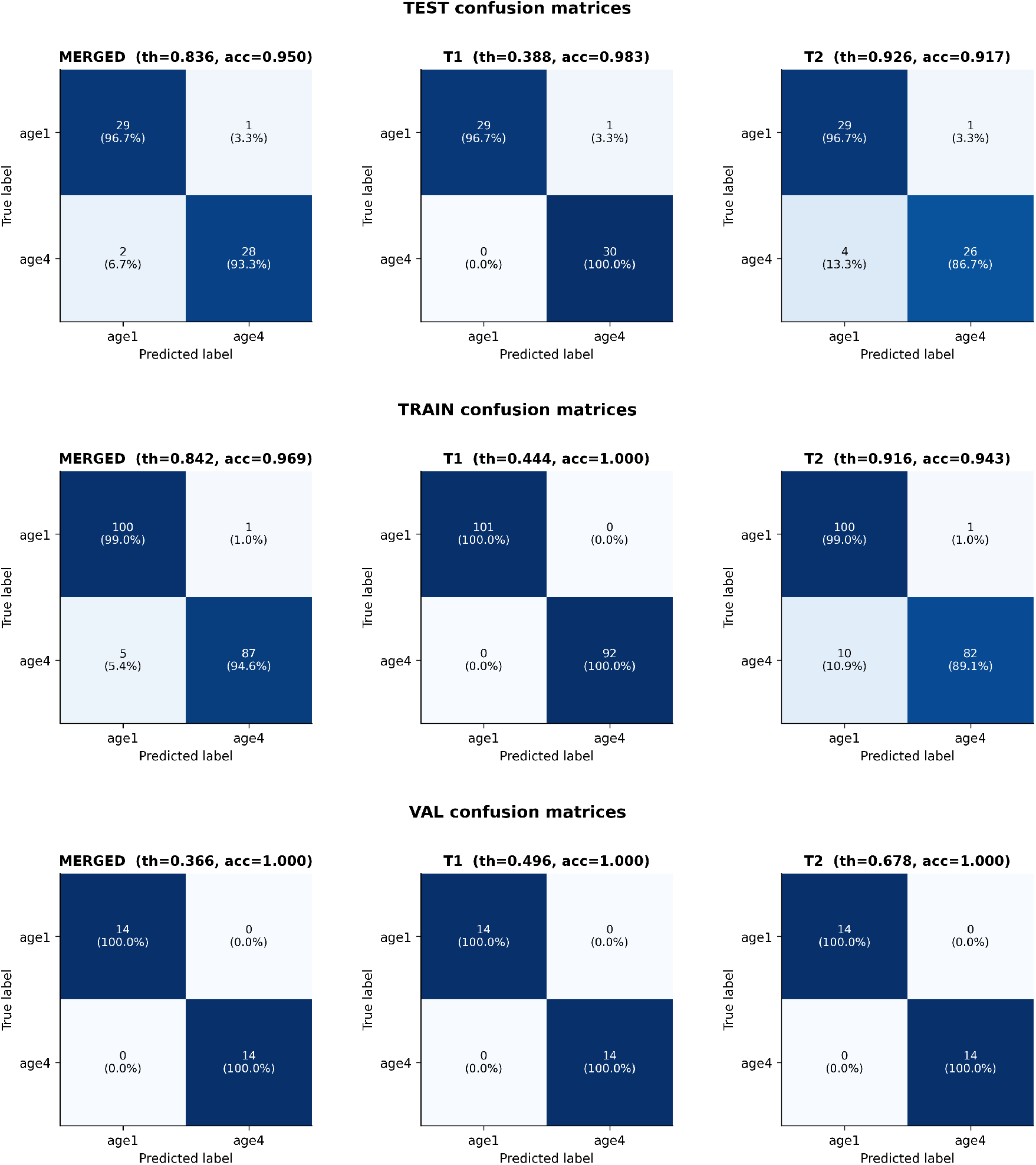
Confusion matrices for TEST (top), TRAIN (middle), and VAL (bottom) sets. Each row shows MERGED, T1-only, and T2-only results.

The best-performing modality was T1, achieving the highest accuracy in both training and test domains, with a maximum training accuracy of 1.00 and a test accuracy of 0.98. This demonstrates both strong expressiveness and robustness of the model. In contrast, the T2 modality exhibited lower accuracy (0.943 for training and 0.916 for testing), indicating slight underfitting compared with the T1 modality. These results suggest that, for this model, the information contained in the T1 modality is sufficient for the classification task, while the T2 modality may introduce additional noise. Indeed, the combined posterior from T1+T2 achieved a training accuracy of 0.969 and a test accuracy of 0.95, which was slightly worse than using T1 alone. The distributions of the posterior probabilities for the *age1* class are shown in Figure 9.

**Figure 9.**
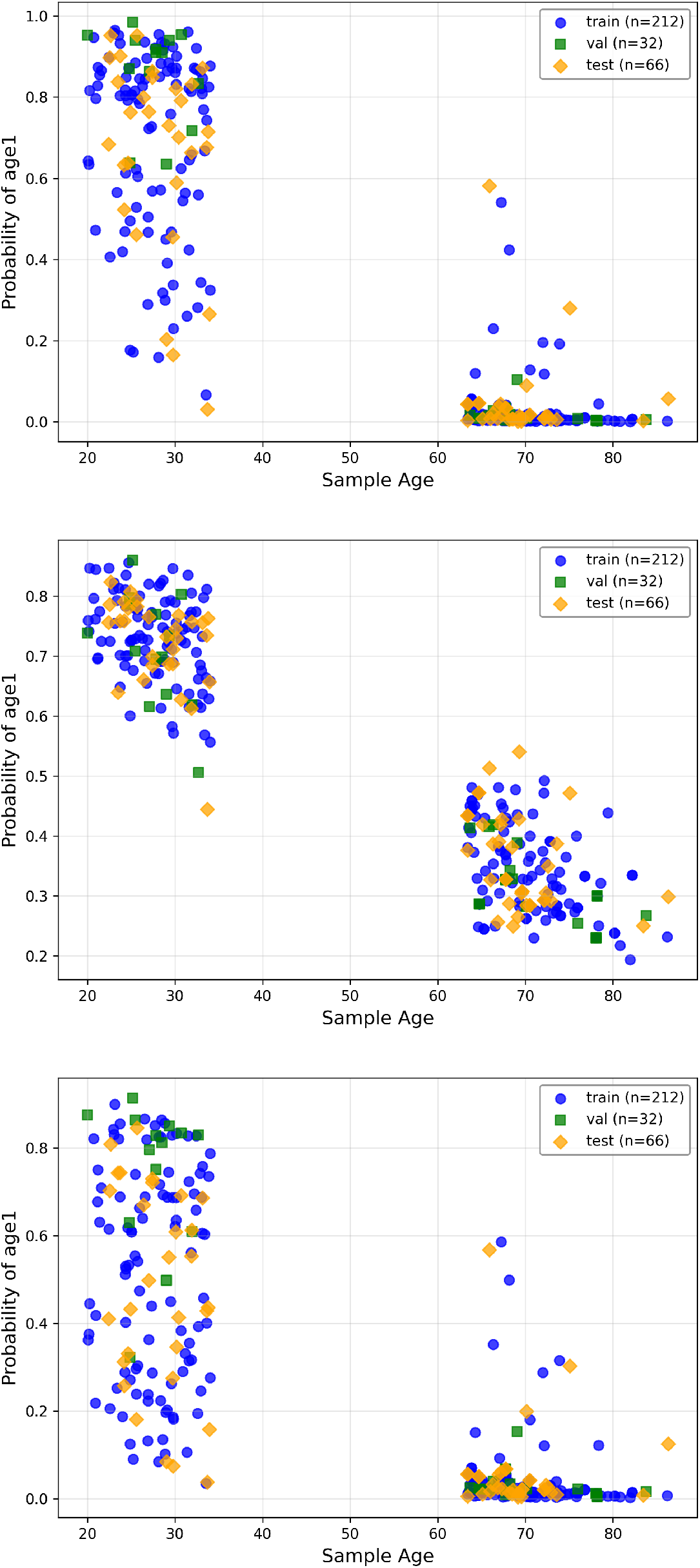
Scatter plots of posterior probability *P* (age1) versus chronological age for MERGED (top), T1-only (middle), and T2-only (bottom).

### B.2 Patch-level Probabilistic Attribution for 3D multimodal image classification

As described in Section 2.5, our model can visualize patch-level contributions to the classification decision even in the 3D multimodal setting. To demonstrate this capability, we visualized the contribution score of every patch across all slices of the input MRI volume. One example of the resulting heatmaps are shown in Figures 10–13.

**Figure 10.**
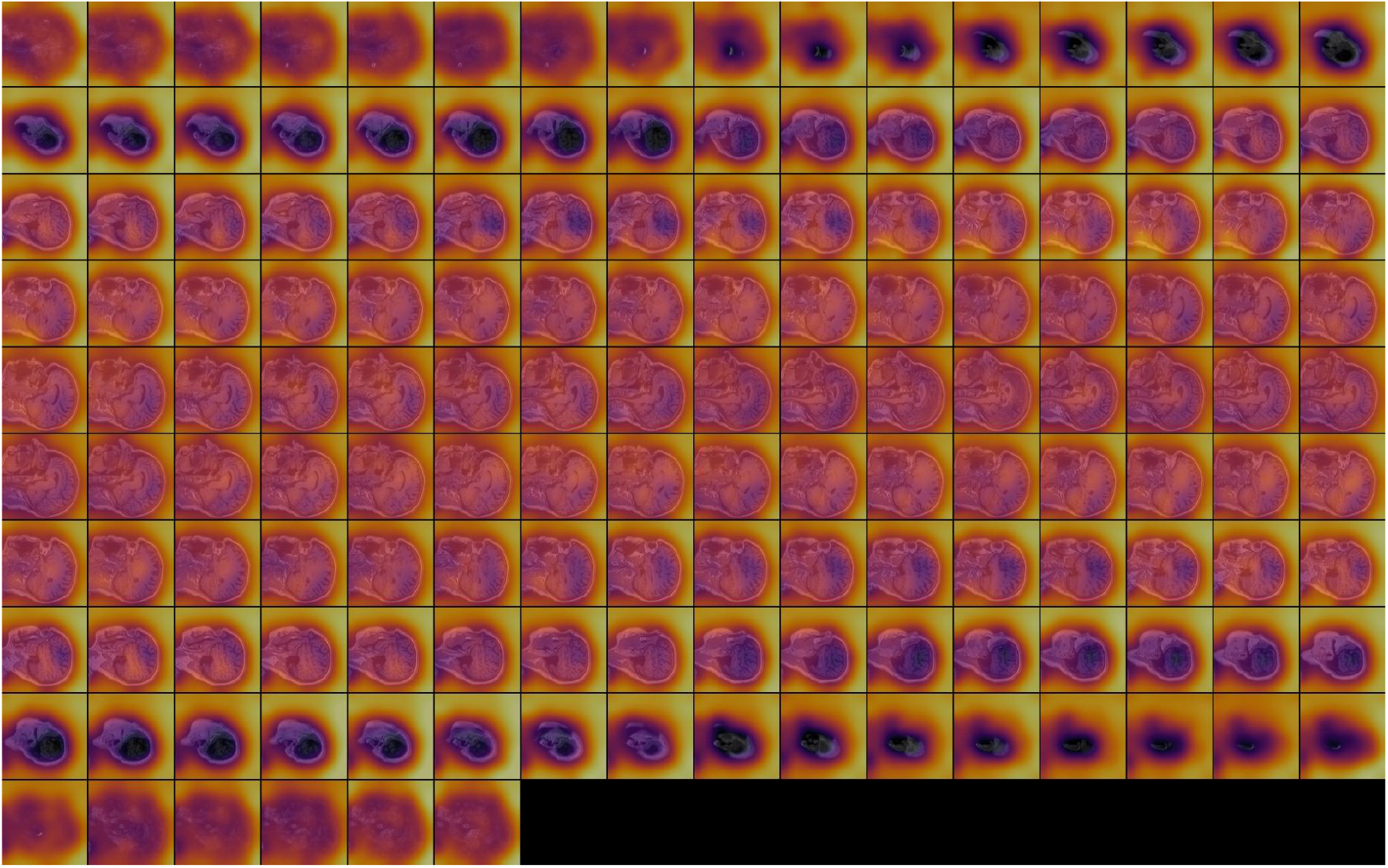
Patch-level contribution heatmap for a sample (true label: age group4 (age range 60-83)). Visualization generated from the T1 modality for the target class **age1**.

**Figure 11.**
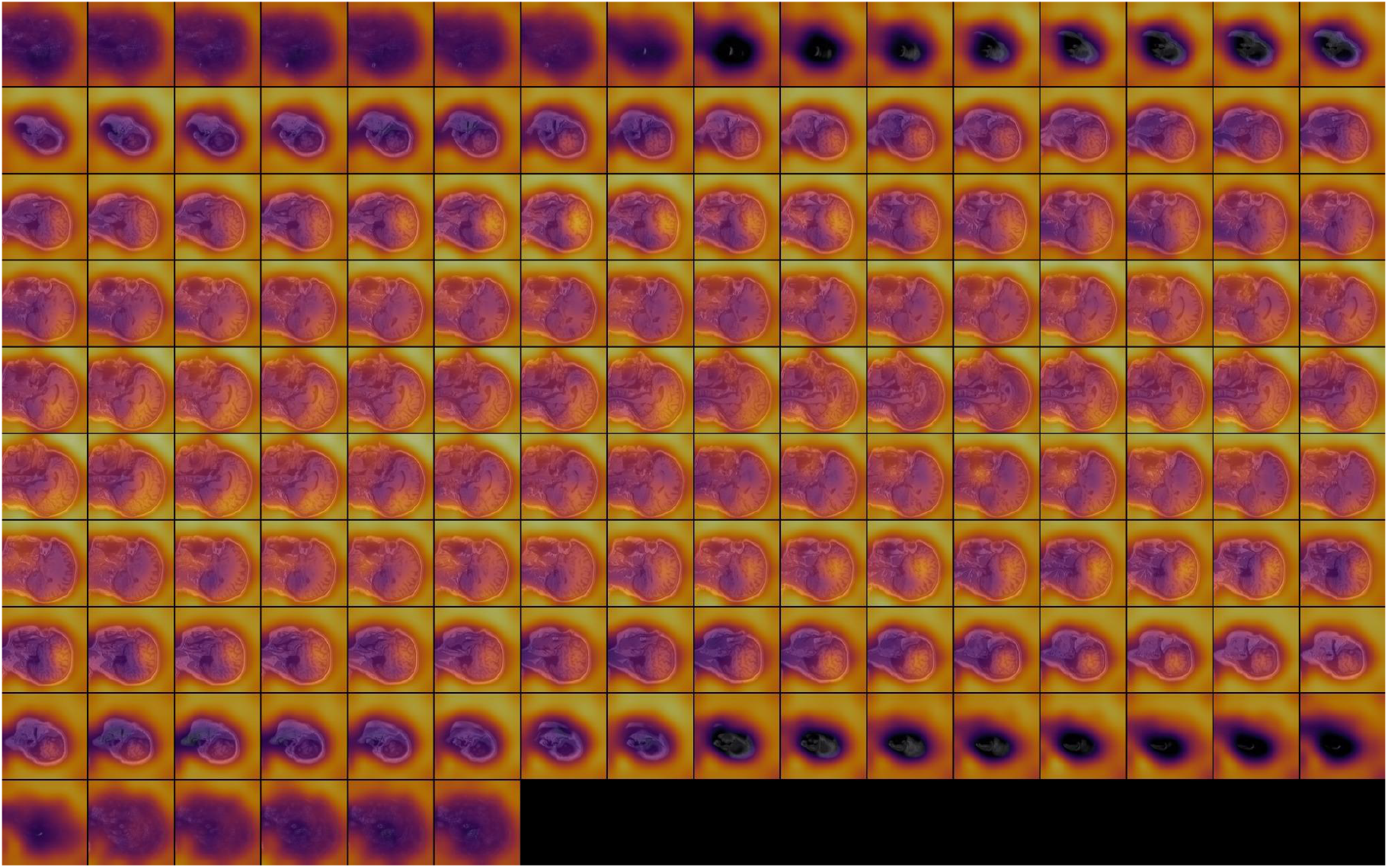
Patch-level contribution heatmap for a sample (true label: age group4 (age range 60-83)). Visualization generated from the T1 modality for the target class **age4**.

**Figure 12.**
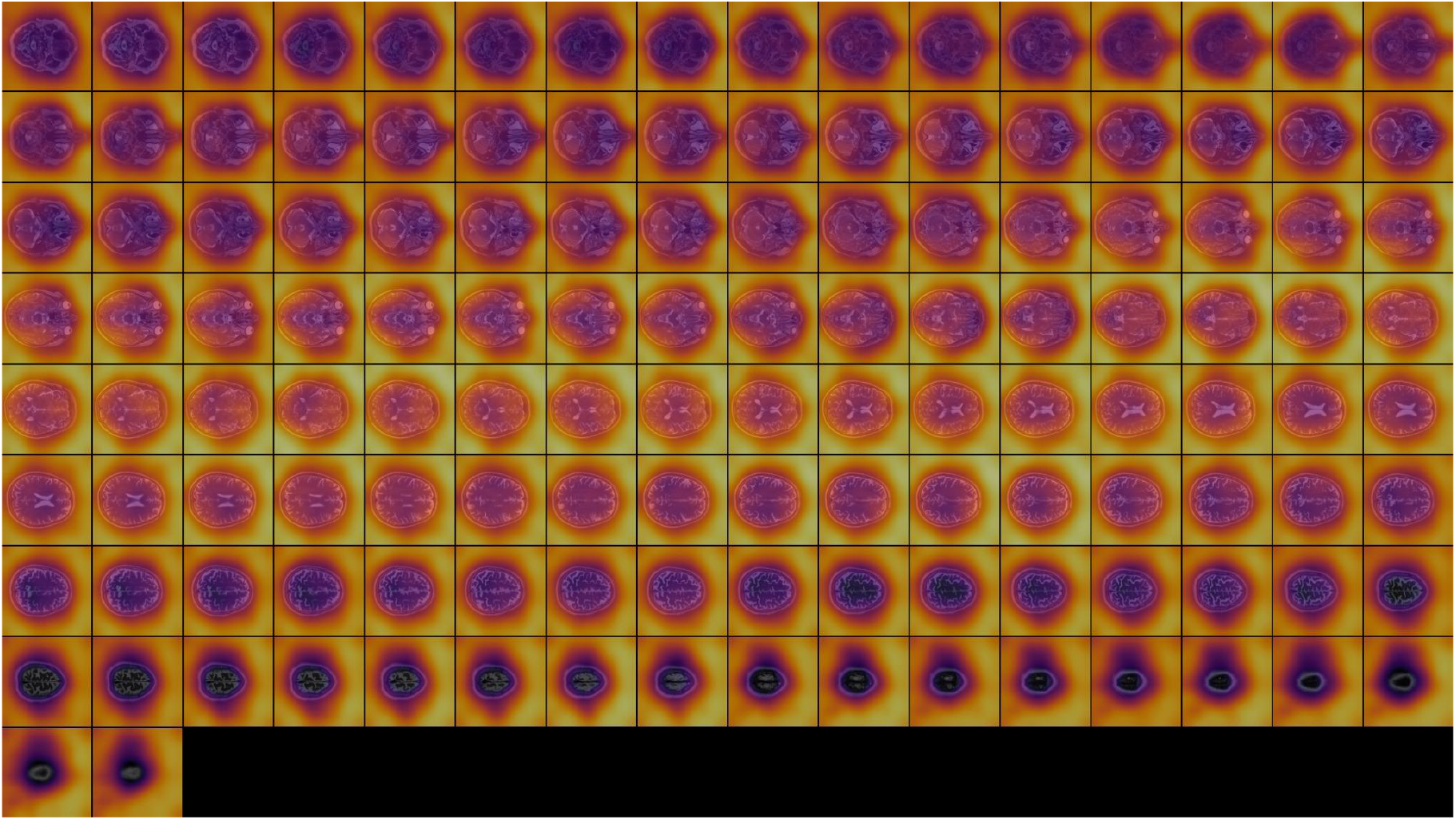
Patch-level contribution heatmap for a sample ((true label: age group4 (age range 60-83)). Visualization generated from the T2 modality for the target class **age1**.

**Figure 13.**
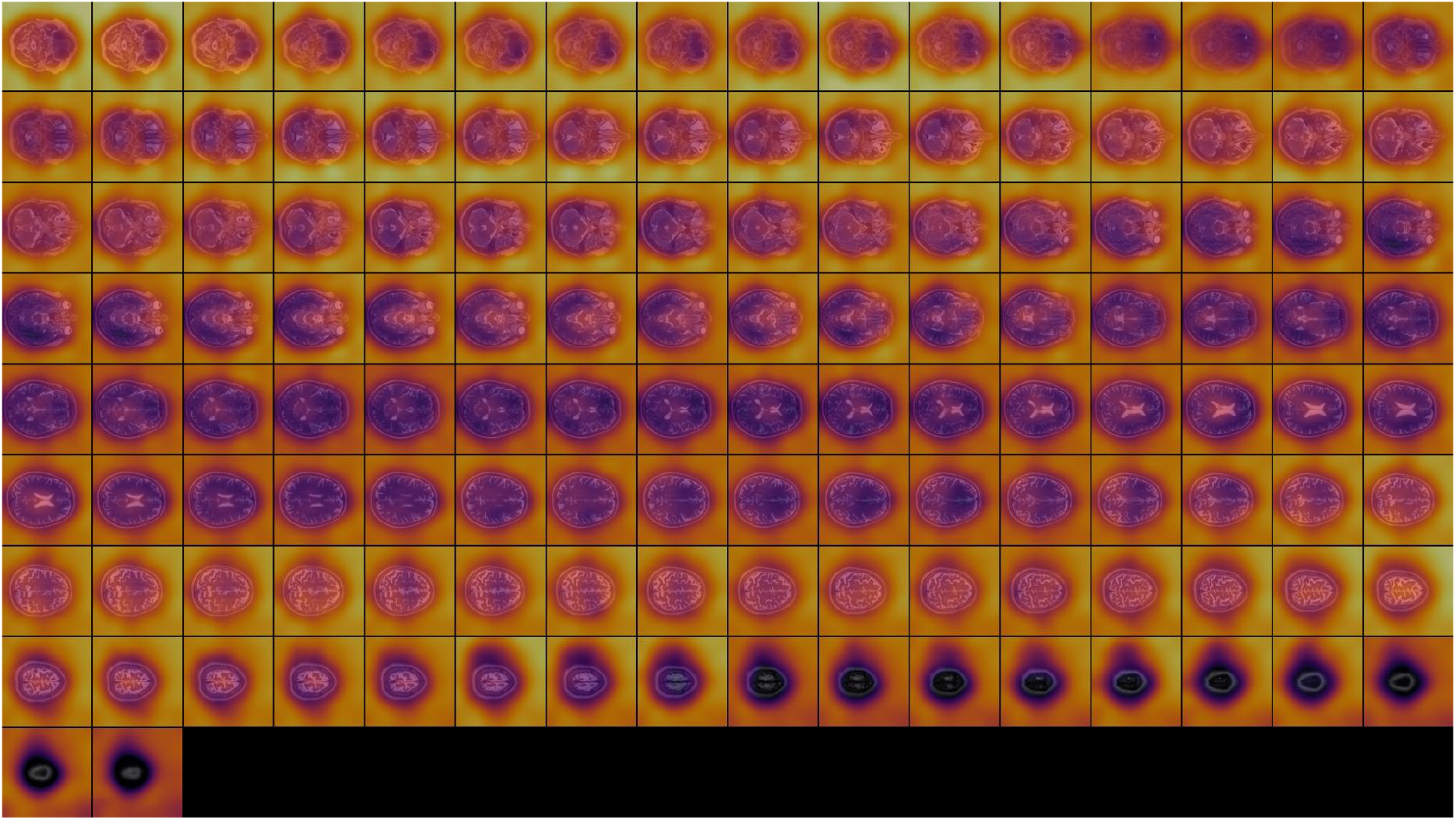
Patch-level contribution heatmap for a sample ((true label: age group4 (age range 60-83)). Visualization generated from the T2 modality for the target class **age4**.

## References

[1] A. Krizhevsky, I. Sutskever, and G. E. Hinton, “Imagenet classification with deep convolutional neural networks,” Advances in neural information processing systems, vol. 25, 2012.

[2] A. Vaswani, N. Shazeer, N. Parmar, J. Uszkoreit, L. Jones, A. N. Gomez, L . Kaiser, and I. Polo-sukhin, “Attention is all you need,” Advances in neural information processing systems, vol. 30, 2017.

[3] C. Patrício, J. C. Neves, and L. F. Teixeira, “Explainable deep learning methods in medical image classification: A survey,” ACM Computing Surveys, vol. 56, no. 4, pp. 1–41, 2023.

[4] R. R. Selvaraju, M. Cogswell, A. Das, R. Vedantam, D. Parikh, and D. Batra, “Grad-cam: Visual explanations from deep networks via gradient-based localization,” in Proceedings of the IEEE international conference on computer vision, pp. 618–626, 2017.

[5] S. Bach, A. Binder, G. Montavon, F. Klauschen, K.-R. Müller, and W. Samek, “On pixel-wise explanations for non-linear classifier decisions by layer-wise relevance propagation,” PloS one, vol. 10, no. 7, p. e0130140, 2015.

[6] A. Dosovitskiy, L. Beyer, A. Kolesnikov, D. Weissenborn, X. Zhai, T. Unterthiner, M. Dehghani, M. Minderer, G. Heigold, S. Gelly, J. Uszkoreit, and N. Houlsby, “An image is worth 16×16 words: Transformers for image recognition at scale,” 2020. arXiv preprint arXiv:2010.11929.

[7] M. T. Ribeiro, S. Singh, and C. Guestrin, ““why should i trust you?” explaining the predictions of any classifier,” in Proceedings of the 22nd ACM SIGKDD international conference on knowledge discovery and data mining, pp. 1135–1144, 2016.

[8] S. M. Lundberg and S.-I. Lee, “A unified approach to interpreting model predictions,” Advances in neural information processing systems, vol. 30, 2017.

[9] S. Jain and B. C. Wallace, “Attention is not explanation,” in Proceedings of the 2019 Conference of the North American Chapter of the Association for Computational Linguistics: Human Language Technologies, Volume 1 (Long and Short Papers), pp. 3543–3556, 2019.

[10] J. Adebayo, J. Gilmer, M. Muelly, I. Goodfellow, M. Hardt, and B. Kim, “Sanity checks for saliency maps,” Advances in neural information processing systems, vol. 31, 2018.

[11] I. E. Kumar, S. Venkatasubramanian, C. Scheidegger, and S. Friedler, “Problems with shapleyvalue-based explanations as feature importance measures,” in International conference on machine learning, pp. 5491–5500, PMLR, 2020.

[12] D. Slack, S. Hilgard, E. Jia, S. Singh, and H. Lakkaraju, “Fooling lime and shap: Adversarial attacks on post hoc explanation methods,” in Proceedings of the AAAI/ACM Conference on AI, Ethics, and Society, pp. 180–186, 2020.

[13] Y. Choi, A. Vergari, and G. Van den Broeck, “Probabilistic circuits: A unifying framework for tractable probabilistic models.” http://starai.cs.ucla.edu/papers/ProbCirc20.pdf, 2020. UCLA Technical Report.

[14] S. Braun, S. Sidheekh, S. Natarajan, A. Vergari, M. Mundt, and K. Kersting, “Autoencoding probabilistic circuits,” in Eighth Workshop on Tractable Probabilistic Modeling, 2025.

[15] W. Chen, S. Yu, H. Shao, L. Sha, and H. Zhao, “Neural probabilistic circuits: An overview,” in Eighth Workshop on Tractable Probabilistic Modeling, 2024.

[16] A. Ciotinga and Y. Choi, “Optimal transport for probabilistic circuits,” arXiv preprint arXiv:2410.13061, 2024.

[17] A. Radford, J. W. Kim, C. Hallacy, A. Ramesh, G. Goh, S. Agarwal, G. Sastry, A. Askell, P. Mishkin, J. Clark, G. Krueger, and I. Sutskever, “Learning transferable visual models from natural language supervision,” in Proceedings of the 38th International Conference on Machine Learning (ICML), 2021. arXiv preprint arXiv:2103.00020.

[18] M. Caron, H. Touvron, I. Misra, H. Jégou, J. Mairal, P. Bojanowski, and A. Joulin, “Emerging properties in self-supervised vision transformers,” in Proceedings of the IEEE/CVF International Conference on Computer Vision (ICCV), 2021. arXiv preprint arXiv:2104.14294.

[19] A. Fateh, Y. Rezvani, S. Moayedi, S. Rezvani, F. Fateh, M. Fateh, and V. Abolghasemi, “Brisc: Annotated dataset for brain tumor segmentation and classification with swin-hafnet,” arXiv preprint arXiv:2506.14318, 2025.

[20] X. Wang, Y. Peng, L. Lu, Z. Lu, M. Bagheri, and R. M. Summers, “Chestx-ray8: Hospital-scale chest x-ray database and benchmarks on weakly-supervised classification and localization of common thorax diseases,” in Proceedings of the IEEE conference on computer vision and pattern recognition, pp. 2097–2106, 2017.

[21] N. Codella, V. Rotemberg, P. Tschandl, M. E. Celebi, S. Dusza, D. Gutman, B. Helba,A. Kalloo, K. Liopyris, M. Marchetti, et al., “Skin lesion analysis toward melanoma detection 2018: A challenge hosted by the international skin imaging collaboration (isic),” arXiv preprint arXiv:1902.03368, 2019.

[22] P. Tschandl, C. Rosendahl, and H. Kittler, “The ham10000 dataset: a large collection of multi-source dermatoscopic images of common pigmented skin lesions,” Scientific Data, vol. 5, p. 180161, 2018.

[23] P. Bergmann, K. Batzner, M. Fauser, D. Sattlegger, and C. Steger, “The mvtec anomaly detection dataset: A comprehensive real-world dataset for unsupervised anomaly detection,” International Journal of Computer Vision, vol. 129, no. 4, pp. 1038–1059, 2021.

[24] R. Peharz, A. Vergari, K. Stelzner, A. Molina, X. Shao, M. Trapp, K. Kersting, and Z. Ghahramani, “Random sum-product networks: A simple and effective approach to probabilistic deep learning,” in Uncertainty in Artificial Intelligence, pp. 334–344, PMLR, 2020.

[25] C. E. Shannon, “A mathematical theory of communication,” Bell System Technical Journal, vol. 27, no. 3, pp. 379–423, 1948.

[26] A. Liu and G. Van den Broeck, “Tractable regularization of probabilistic circuits,” Advances in Neural Information Processing Systems, vol. 34, pp. 3558–3570, 2021.

[27] A. Bhattacharya, D. Pati, and Y. Yang, “Bayesian fractional posteriors,” 2019.

[28] D. P. Kingma and J. Ba, “Adam: A method for stochastic optimization,” International Conference on Learning Representations (ICLR), 2015.

[29] K. Moreland, “Diverging color maps for scientific visualization,” in Advances in Visual Computing (ISVC), vol. 5876 of Lecture Notes in Computer Science, pp. 92–103, Springer, 2009.

[30] L. Arras, G. Montavon, K.-R. Müller, and W. Samek, “Explaining recurrent neural network pre-dictions in sentiment analysis,” in Proceedings of the 8th Workshop on Computational Approaches to Subjectivity, Sentiment and Social Media Analysis, pp. 159–168, 2017.

[31] M. Bohle, F. Eitel, M. Weygandt, and K. Ritter, “Layer-wise relevance propagation for explaining deep neural network decisions in mri-based alzheimer’s disease classification,” Frontiers in Aging Neuroscience, vol. 11, p. 194, 2019.

[32] M. Komorowski et al., “Towards evaluating explanations of vision transformers for medical imaging,” in Proceedings of the IEEE/CVF Conference on Computer Vision and Pattern Recognition Workshops (CVPRW), pp. 6133–6142, 2023.

[33] A. Rao, J. Park, S. Woo, J.-Y. Lee, and O. Aalami, “Studying the effects of self-attention for medical image analysis,” in Proceedings of the IEEE/CVF International Conference on Computer Vision, pp. 3416–3425, 2021.

[34] T. Hastie and R. Tibshirani, “Generalized additive models,” Statistical science, vol. 1, no. 3, pp. 297–310, 1986.

[35] H. Nori, S. Jenkins, P. Koch, and R. Caruana, “Interpretml: A unified framework for machine learning interpretability,” arXiv preprint arXiv:1909.09223, 2019.

[36] R. Agarwal, L. Melnick, N. Frosst, X. Zhang, B. Lengerich, R. Caruana, and G. E. Hinton, “Neural additive models: Interpretable machine learning with neural nets,” Advances in neural information processing systems, vol. 34, pp. 4699–4711, 2021.

[37] T. Kusumoto, “Dvpnet: A new xai-based interpretable genetic profiling framework using nucleotide transformer and probabilistic circuits,” bioRxiv, pp. 2026–01, 2026.

[38] H. Dalla-Torre, L. Gonzalez, J. Mendoza-Revilla, N. Lopez Carranza, A. H. Grzywaczewski, F. Oteri, C. Dallago, E. Trop, B. P. De Almeida, H. Sirelkhatim, et al., “Nucleotide transformer: building and evaluating robust foundation models for human genomics,” Nature Methods, vol. 22, no. 2, pp. 287–297, 2025.

[39] A. Liu, K. Ahmed, and G. V. d. Broeck, “Scaling tractable probabilistic circuits: A systems perspective,” arXiv preprint arXiv:2406.00766, 2024.

[40] IXI – Information eXtraction from Images Project, “Ixi dataset.” https://brain-development.org/ixi-dataset/, 2006. Accessed: 2025-11-12.

